# Hospitalized patients with severe COVID-19 during the Omicron wave in Israel - benefits of a fourth vaccine dose

**DOI:** 10.1101/2022.04.24.22274237

**Authors:** Tal Brosh-Nissimov, Khetam Hussein, Yonit Wiener-Well, Efrat Orenbuch-Harroch, Meital Elbaz, Shelly Lipman-Arens, Yasmin Maor, Yael Yagel, Bibiana Chazan, Mirit Hershman-Sarafov, Galia Rahav, Oren Zimhony, Adi Zaidman Shimshovitz, Michal Chowers

**Affiliations:** Faculty of Health Sciences, Ben Gurion University of the Negev, Beer Sheba, Israel; Infectious Diseases Unit, Samson Assuta Ashdod University Hospital, Ashdod, Israel; Rambam Health care campus, Haifa, Israel; Rappaport Faculty of Medicine, Technion-Israel Institute of Technology, Haifa, Israel; Shaare Zedek Medical Center, Jerusalem, Israel; Faculty of Medicine, Hebrew University of Jerusalem, Jerusalem, Israel; Division of Microbiology and Infectious Diseases, Hadassah Hebrew University Medical Center, Jerusalem, Israel; Department of Infectious Diseases, Tel Aviv Sourasky Medical Center; Sackler Faculty of Medicine, Tel Aviv University, Tel Aviv, Israel; Infectious disease and Infection Control Unit, Hillel Yaffe Medical Center, Hadera, Israel; Infectious Disease Unit, Wolfson Medical Center, Holon, Israel; Infectious Disease Institute, Soroka Medical Center, Beer Sheba, Israel; Infectious Diseases Unit, Emek Medical Center, Afula, Israel; Bnai Zion medical center, Haifa, Israel; Infectious Diseases Unit, Sheba Medical Center, Tel Hashomer, Israel; Infectious Diseases Unit, Kaplan Medical Center, Rhovot, Israel; Infectious Disease Unit, The Baruch Padeh Medical Center, Tiberias, Israel; Meir Medical Center, Kfar Saba, Israel

**Keywords:** COVID-19, severe COVID-19, BNT162b2, vaccine, booster, outcome, fourth dose, second booster

## Abstract

**Importance:** Waning immunity against COVID-19 in parallel with an increased incidence during the Omicron outbreak led the Israeli Ministry of Health to recommend a second booster dose of BNT162b2 (Pfizer) to high-risk individuals. Israel was the first country to recommend this, allowing evaluation of the added protection of a fourth vaccine dose to hospitalized patients with severe diseases.

**Objective:** To assess the effect of a fourth dose for hospitalized patients with severe/critical breakthrough COVID-19.

**Design:** A cohort study of hospitalized adults from 01/15/2022–01/31/2022.

**Settings:** A multi-center study of 14 medical centers in Israel.

**Participants:** Hospitalized adult patients with PCR-confirmed severe/critical COVID-19. Excluded were patients lacking data on vaccination status.

**Exposure:** Cases were divided according to the total number of vaccine doses received up to 7 days before diagnosis. Unvaccinated adults and single-dose recipients were grouped into an unvaccinated group.

**Main Outcome:** A composite of mechanical-ventilation or in-hospital death was defined as poor outcome. Outcomes were compared between 3- and 4-dose vaccinees.

**Results:** Included were 1,049 patients with severe/critical COVID-19, median age 80 (IQR 69-87), 51% males. Among them, 360 unvaccinated, 34, 172, 386 and 88 were after 1, 2, 3 or 4 doses, respectively. Patients after 3 doses were older, had more males and immunosuppression, but with similar outcomes, 49% vs. 51% compared to unvaccinated patients (p=0.72). Patients after 4 doses were similarly older and immunosuppressed, but had improved outcomes compared to unvaccinated patients, 34% vs. 51% (p<0.01). We proceeded to examine independent predictors for poor outcome in fully-vaccinated patients with either 3 doses given a median of 161 (IQR 147-168) days earlier, or 4 doses given a median of 14 (IQR 10-18) days before diagnoses. Receipt of the fourth dose conferred significant protection: OR 0.51 (95%CI 0.30.87).

**Conclusion and Relevance:** Within a population of hospitalized patients with severe/critical breakthrough COVID-19, a recent fourth dose was associated with significant protection against mechanical ventilation or death, compared to fully vaccinated single-boosted individuals.

**Key points:** *Question:* What is the benefit of a fourth vaccine dose (second booster) for hospitalized patients with severe COVID-19?

*Findings:* In this multicenter cohort study in Israel during the Omicron wave, hospitalized severe COVID-19 patients that received a recent fourth dose had a 49% lower odds for a poor outcome (mechanical ventilation or death) compared with those who received 3 doses approximately 5 months before diagnosis, a significant difference.

*Meaning:* A vaccine booster given at the onset of a COVID-19 wave can benefit vulnerable individuals.

## Introduction

Vaccination against SARS-CoV-2 has resulted in a significant change in the response to COVID-19 since the end of 2021. Nevertheless, due to waning immunity [1–3], a third dose given as a booster became an essential countermeasure during the Delta variant wave a few months later [4,5]. The spread of the Omicron variant has challenged even the most vaccinated populations, with lower vaccine effectiveness (VE) and high rate of breakthrough infections [6–8]. On January 2, 2022, the Israeli Ministry of Health recommended a fourth dose (second booster) for individuals age 60 years and older and immunocompromised patients, 4 months after the third dose, anticipating a benefit in the prevention of severe outcomes. Since then, several population studies have shown its benefit in preventing severe COVID-19, hospitalization and death [9–11].

This study assessed the benefit of a fourth vaccine dose, compared to three doses, for hospitalized patients with severe or critical breakthrough COVID-19.

## Methods

This multi-center cohort study included adult patients hospitalized in 14 participating hospitals due to severe or critical COVID-19. Electronic medical records of adult patients reported to have PCR-confirmed severe or critical COVID-19 during their stay were reviewed by an infectious disease specialist. COVID-19 severity was defined according to the National Institute of Health guidelines [12]. Patients without valid data regarding previous vaccinations or lacking clinical data, and patients who did not have severe/critical COVID-19 upon retrospective case review were excluded. Cases were divided into cohorts according to the number of vaccine doses received at least 7 days prior to diagnosis. The vaccine type was not recorded, but almost all vaccines given in Israel were BNT162b2 (Pfizer). The primary composite outcome of the study was mechanical ventilation (MV) or in-hospital death, referred to as poor outcome. For inter-group comparisons, patients who received no or only one dose were considered unvaccinated, were grouped together and compared separately to vaccinated patients who received three or four doses.

To assess the benefit of the fourth dose for the 3-dose boosted population, we performed an outcome analysis on the entire group of vaccinated patients (3 or 4 doses) with the number of doses as one of the independent variables.

When available, we recorded the results of SARS-CoV-2 RNA sequencing. Nevertheless, the national sequencing data showed that the most common circulating variant during the study period was Omicron, constituting 90-99% of sequenced isolates [13].

### Statistical analysis

Variables were compared between vaccinated groups and between patients with a good or a poor outcome. Categorical variables were compared using chi-square or Fisher’s exact tests, and continuous variables were compared using Mann-Whitney test. Multivariate analysis of risk factors for poor outcome was performed with logistic regression on clinically meaningful variables, and variables with p<0.1 on univariant analysis with the enter method. All tests were two-tailed. IBM SPSS-25 was used for all analyses.

### Ethics approval

The study was approved by the Institutional Research Ethics Boards of each participating hospital, and overall by the Assuta-Ashdod Hospital board (#0027-22-AAA). Due to the retrospective design, informed consent was not required.

## Results

From national data during the 2-week study period, by January 15, 2022, 487,211 Israelis age 60 years or older had received a fourth dose of vaccine, constituting 31% of the eligible population. Another 108,643 (7%) individuals received a fourth dose by the end of the study, January 31, 2022. During the study period, 2,602 patients were hospitalized in Israel with severe-critical COVID-19. Among them, 862 were unvaccinated, and 106, 393, 947 and 294 had received 1, 2, 3 or 4 doses, respectively, at least 7 days before their admission (source: R. Singer, Ministry of Health, personal communication).

From January 15, 2022 to January 31, 2022, 1,237 cases reported with severe/critical COVID-19 in the participating hospitals were reviewed. After excluding 188 patients, 1,049 patients with verified severe COVID-19 with a known vaccination history were analyzed (Figure 1). These constituted 40% of the nationally reported cases during the study period. Groups included 360 (42% of national data) unvaccinated patients, 43 (41%) after 1 dose, 172 (44%) after 2 doses, 386 (41%) after 3 and 88 (30%) after 4 doses. The median age was 80 (IQR 69-87) years, 535 (51%) were males, and 28 (2.7%) had history of previous COVID-19. Among the 138 (13%) patients with a viral RNA sequencing result, all were Omicron variant.

**Figure 1.**
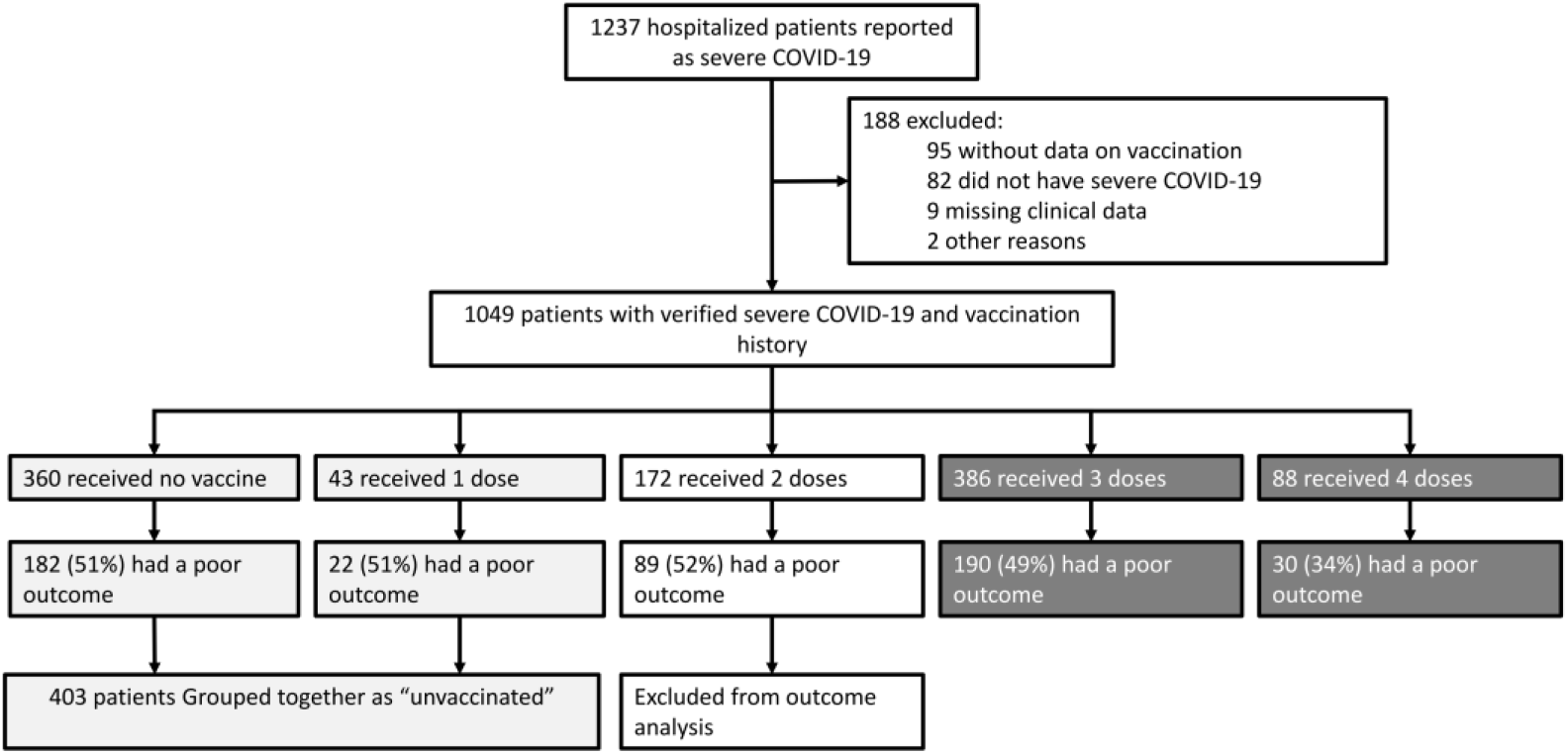
Study population Flowchart displaying the inclusion of patients and selection of cohorts according to vaccination status. Light-grey boxes contain the unvaccinated groups (no or 1 vaccine dose). Dark-grey boxes contain the fully-vaccinated groups (3 or 4 doses).

We compared unvaccinated patients (no vaccination or only one dose) to fully-vaccinated patients with 3 or 4 doses. The 172 patients who received only 2 doses were not included in the analysis, as they were deemed to have partial immune status, with a median time from the second dose of 326 days (IQR 255-360). The 3-dose group patients received their third dose a median of 161 days (IQR 147-168), while the 4-dose group patients received the fourth dose a median of 14 days (IQR 10-18) before admission (p<0.01).

Comparison of unvaccinated patients to those with 3 doses is shown in Table 1. The 3-dose vaccinees were older, had a higher frequency of long-term care facility residence, hypertension, chronic renal failure (CRF), cancer, immunosuppression, lower rate of previous COVID-19, and received fewer treatments with baricitinib and more convalescent plasma therapy. The rates of death and MV were similar between groups (49% vs. 51%, p=0.72).

**Table 1:** Comparison between unvaccinated and 3 or 4-dose vaccinated hospitalized patients with severe-critical COVID-19

| Variable | Unvaccinated<br>(0/1 doses) | Received 3<br>doses | P-value<br>(0 vs. 3) | Received 4<br>doses | P-value<br>(0 vs. 4) |
| --- | --- | --- | --- | --- | --- |
| N | 403 | 386 |  | 88 |  |
| Time from last vaccine dose,<br>days (IQR) | NA | 161 (147-168) | NA | 14 (10-18) | NA |
| Age, median (IQR) | 78 (67-86) | 81 (70-88) | **0.03** | 83 (74-88) | **<0.01** |
| Male sex, n (%) | 190 (47%) | 206 (53%) | 0.09 | 49 (56%) | 0.16 |
| Long term care facility<br>residence, n (%) | 53 (13%) | 73 (19%) | **0.03** | 34 (39%) | **<0.01** |
| **Comorbidities, n (%)** |  |  |  |  |  |
| Diabetes mellitus | 178 (45%) | 178 (46%) | 0.67 | 38 (43%) | 0.91 |
| Hypertension | 261 (65%) | 278 (72%) | **0.03** | 63 (72%) | 0.26 |
| BMI >30 | 110/358 (31%) | 78/335 (23%) | **0.03** | 18/74 (24%) | 0.33 |
| Ischemic heart disease | 124 (31%) | 122 (32%) | 0.82 | 30 (34%) | 0.61 |
| Congestive heart failure | 125 (31%) | 125 (32%) | 0.7 | 20 (23%) | 0.16 |
| Chronic renal failure | 99 (25%) | 122 (32%) | **0.03** | 26 (30%) | 0.34 |
| Chronic liver disease | 15 (4%) | 9 (2%) | 0.3 | 2 (2%) | 0.56 |
| Chronic lung disease | 94 (23%) | 96 (25%) | 0.62 | 25 (29%) | 0.34 |
| Dementia | 135 (34%) | 109 (28%) | 0.12 | 33 (38%) | 0.54 |
| Cancer | 50 (12%) | 79 (21%) | **<0.01** | 20 (23%) | **0.01** |
| Immunosuppression, n (%) | 33 (8%) | 79 (21%) | **<0.01** | 31 (36%) | **<0.01** |
| Past COVID-19 | 15 (4%) | 3 (1%) | **<0.01** | 0 (0%) | 0.09 |
| **Treatment before severe COVID-19** |  |  |  |  |  |
| Nirmaltrevir/ritonavir | 5 (1%) | 8 (2%) | 0.41 | 4 (5%) | 0.06 |
| Molnupiravir | 5 (1%) | 3 (1%) | 0.73 | 2 (2%) | 0.61 |
| **Treatment during admission** |  |  |  |  |  |
| Oxygen | 388 (96%) | 376 (97%) | 0.42 | 85 (97%) | 1.0 |
| High flow nasal canula | 148 (37%) | 130 (35%) | 0.5 | 24 (27%) | 0.11 |
| Mechanical ventilation | 83 (20%) | 85 (22%) | 0.66 | 14 (16%) | 0.38 |
| ECMO | 7 (2%) | 3 (1%) | 0.34 | 1 (1%) | 1.0 |
| Steroids | 375 (93%) | 357 (93%) | 0.79 | 82 (93%) | 1.0 |
| Remdesivir | 201 (50%) | 182 (47%) | 0.43 | 55 (63%) | **0.04** |
| Anti-IL6 | 21 (5%) | 21 (6%) | 1.0 | 3 (3%) | 0.6 |
| Baricitinib | 44 (11%) | 25 (7%) | **0.03** | 7 (8%) | 0.45 |
| Convalescent plasma | 1 (<1%) | 8 (2%) | **0.02** | 2 (2%) | 0.09 |
| Death | 188 (47%) | 170 (44%) | 0.48 | 26 (30%) | **<0.01** |
| Hospital stay, days, median (IQR) | 6 (4-9) | 6 (3-10) | 0.31 | 5 (3-10) | 0.36 |
| Poor outcome (death or mechanical ventilation) | 204 (51%) | 190 (49%) | 0.72 | 30 (34%) | **<0.01** |

Abbreviations: IQR, interquartile range; ECMO, extracorporeal membrane oxygenation; IL6, interleukin 6.

Next, we compared the unvaccinated patients to the 4-dose vaccinated patients (Table 1): The 4-dose vaccinated cohort were older, more lived in long-term care facilities, were on immunosuppression, and remdesivir treatment. The rate of death or MV was significantly lower for 4-dose patients (34% vs. 51%, p<0.01.

As a significant difference in the primary outcome was shown only for the 4-dose group of vaccinated patients, we evaluated the risk factors for a poor outcome within the fully-vaccinated patient group (3 or 4 doses) in univariate analysis (Table 2), followed by multivariate regression analysis (Figure 2). Receipt of a fourth dose was shown to confer significant protection against a poor outcome compared with three doses, with an odds ratio of 0.51 (95%CI 0.3-0.87). Other variables associated with protection from a poor outcome were chronic lung disease and remdesivir treatment, while male sex, dementia and chronic renal failure were detrimental. Immunosuppression showed a trend for a worse outcome (OR 1.58, 95%CI 0.98-2.54, p=0.06). Age did not influence outcome in this fully vaccinated group.

**Table 2:**
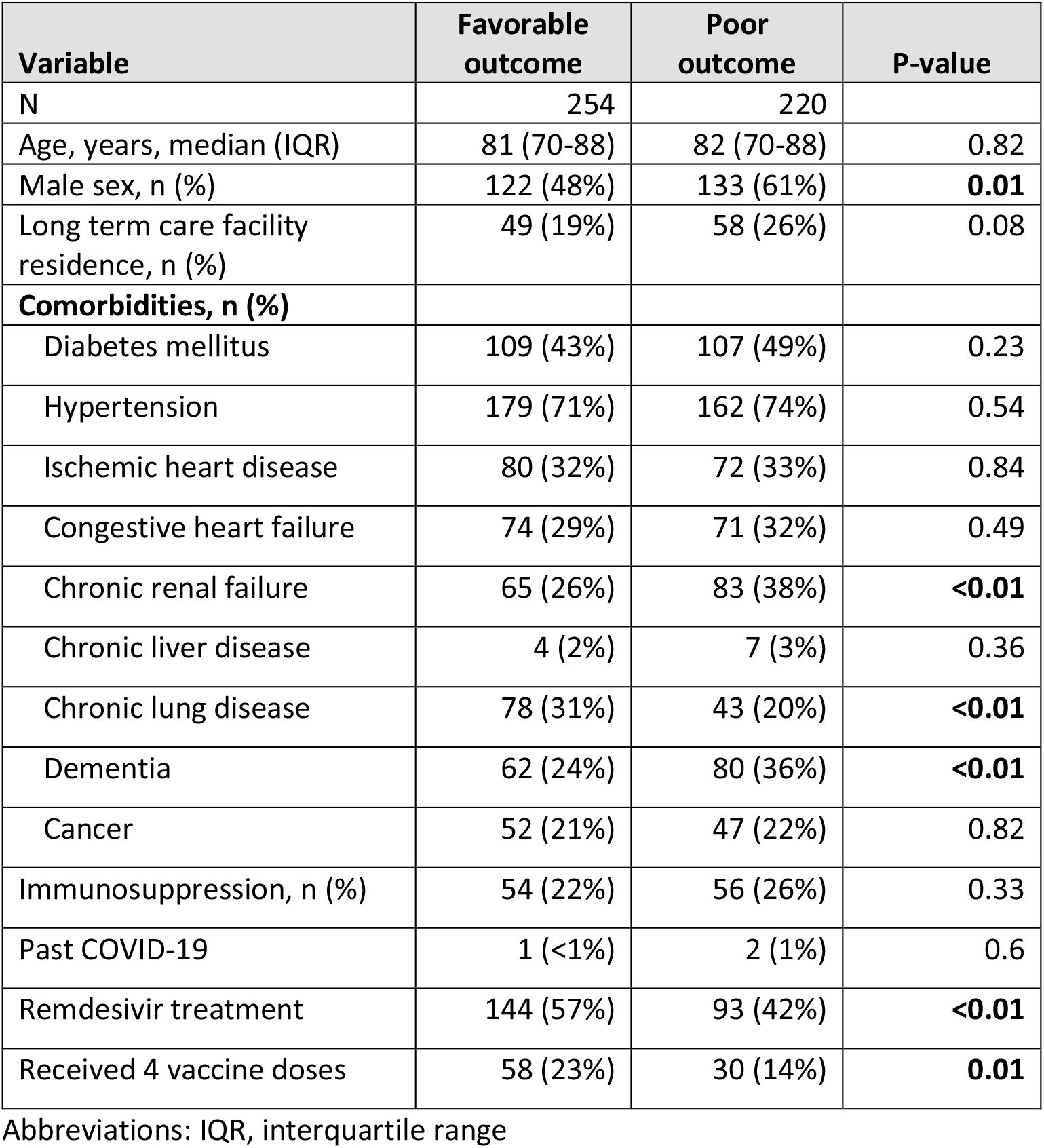
Univariate comparison of patients with a favorable vs. poor outcome, defined as death or mechanical ventilation

| Variable | Favorable outcome | Poor outcome | P-value |
| --- | --- | --- | --- |
| N | 254 | 220 |  |
| Age, years, median (IQR) | 81 (70-88) | 82 (70-88) | 0.82 |
| Male sex, n (%) | 122 (48%) | 133 (61%) | **0.01** |
| Long term care facility residence, n (%) | 49 (19%) | 58 (26%) | 0.08 |
| **Comorbidities, n (%)** |  |  |  |
| Diabetes mellitus | 109 (43%) | 107 (49%) | 0.23 |
| Hypertension | 179 (71%) | 162 (74%) | 0.54 |
| Ischemic heart disease | 80 (32%) | 72 (33%) | 0.84 |
| Congestive heart failure | 74 (29%) | 71 (32%) | 0.49 |
| Chronic renal failure | 65 (26%) | 83 (38%) | **<0.01** |
| Chronic liver disease | 4 (2%) | 7 (3%) | 0.36 |
| Chronic lung disease | 78 (31%) | 43 (20%) | **<0.01** |
| Dementia | 62 (24%) | 80 (36%) | **<0.01** |
| Cancer | 52 (21%) | 47 (22%) | 0.82 |
| Immunosuppression, n (%) | 54 (22%) | 56 (26%) | 0.33 |
| Past COVID-19 | 1 (<1%) | 2 (1%) | 0.6 |
| Remdesivir treatment | 144 (57%) | 93 (42%) | **<0.01** |
| Received 4 vaccine doses | 58 (23%) | 30 (14%) | **0.01** |

Abbreviations: IQR, interquartile range

**Figure 2.**
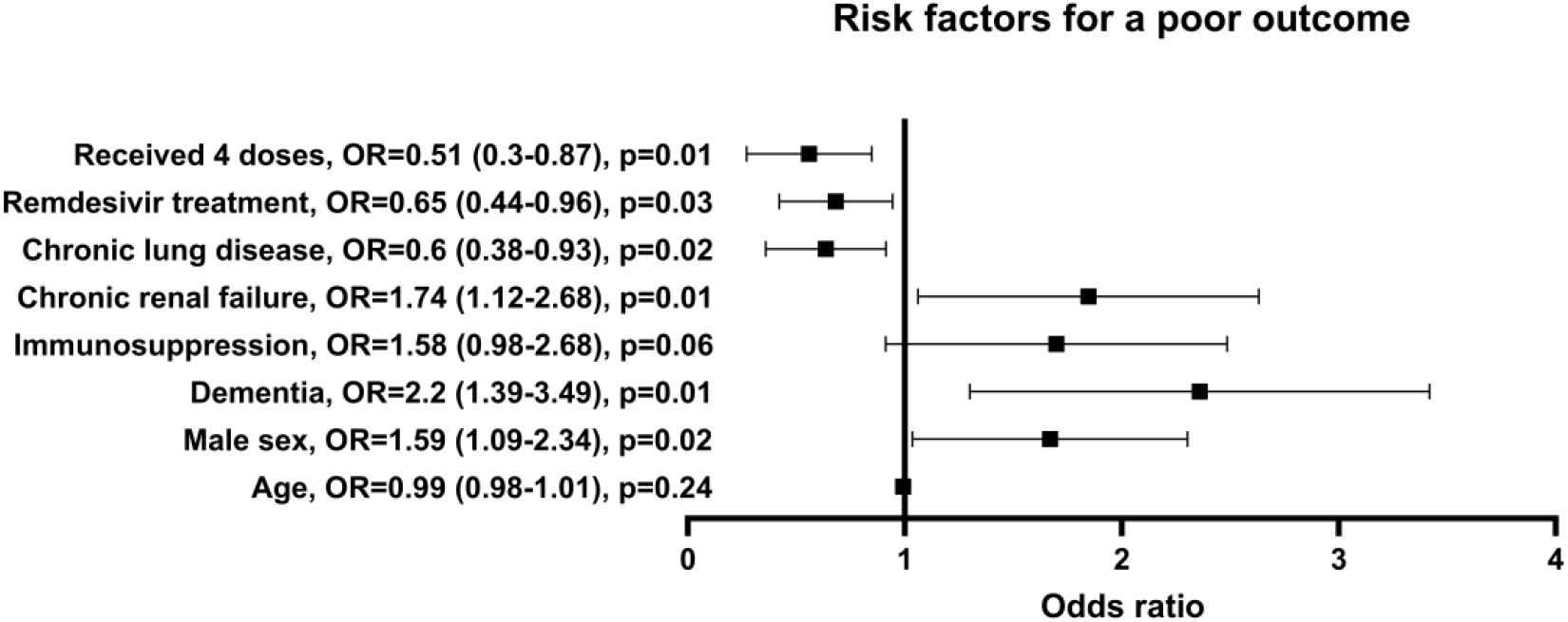
Risk factors for poor outcome within vaccinated patients: Box and whisker plot displaying a regression analysis of risk factors for a poor composite outcome of death or mechanical ventilation, within the population of vaccinated (3 or 4 doses) patients with severe COVID-19. Blocks and whiskers signify odds ratio (poor vs. good outcome) and 95% confidence intervals (CI).

## Discussion

This study analyzed clinical data from 1,049 adult patients with severe/critical COVID-19 who were admitted to 14 general hospitals in Israel in a two-week period in January 2022, during a COVID-19 wave with a predominantly Omicron variant. Fully vaccinated adults with either 3 or 4 vaccines were older, and more were immunocompromised compared to the unvaccinated patients. A fourth vaccine (received a median of two weeks prior) provided significant protection from death or MV (OR 0.51 (95%CI 0.3-0.87)) to its older, immunocompromised patient population, whereas three vaccine doses (last dose received a median of 23 weeks prior) did not provide this protection.

Vaccine effectiveness (VE) against various clinical outcomes was shown to decrease during the Omicron wave, due to the antigenic distance of this variant and waning immunity. VE against symptomatic infection was at best 67.2% shortly after a third BNT162b2 dose and declined to 47.5% after 10 or more weeks [6]. VE against hospitalization after a 3-dose BNT162b2 vaccination schedule decreased from 91% within 2 months of vaccination to 78% beyond 4 months [8]. VE against MV or death was 94% after 3 doses during the Omicron period in another study [7], but the median time from the third dose was only 60 days.

The waning immunity after three doses and the fact that most of the older Israeli population was more than 4-5 months after their third dose at the onset of the Omicron wave, led the Israeli Ministry of Health to recommend a fourth dose to individuals age 60 years or older, those with comorbidities and healthcare personnel on January 2, 2022. Since then, several studies showed high protection afforded by a fourth dose against severe disease and death. Compared with individuals who received 3 doses, those who received a fourth dose had a 3.5-fold lower rate of severe disease during a 6-week follow up, in a national observational study [9]. VE against infection was modest and declined rapidly. Two other studies comparing four- to three-dose recipients, reported VE of 64-73% against severe disease at a 4-9 week follow-up [10,11], and 88% against mortality during a 10-week follow-up [14]. Our findings show another added benefit from the fourth dose. Even after failure of that dose to prevent infection and progression to severe disease, it was associated with greater protection from the most severe outcomes.

In a previous study on breakthrough infections during the Delta wave in Israel, we showed that although vaccinated patients were considerably older and more immunocompromised, poor outcome, once hospitalized, was not different between vaccinated and unvaccinated patients [15]. In that study, the vaccinated cohort included patients who received two doses of BNT162b2 approximately 6 months earlier. Those results echo those of our present study in the sub-group of the patients receiving 3-doses 5 months before infection. This is not to imply that vaccination did not have an effect on disease outcomes. It prevented hospitalization of the younger and healthier population, as can be seen by the differences in age and co-morbidities between the unvaccinated and 3-dose vaccinees. The current study enabled us to compare fully-vaccinated patients with a breakthrough infection from a single variant, with various intervals from their last booster. The data presented here, suggest that the observed benefit of this additional dose might not be due to a specific immunogenicity of a fourth dose, but to its temporal proximity to infection.

Other independent variables associated with protection against poor outcomes were treatment with remdesivir and chronic lung diseases. Improved outcomes with remdesivir were expected [16,17]. More surprising was improved outcomes of patients with chronic lung infection. A possible explanation is low baseline oxygenation, wrongfully diagnosed as severe COVID-19. A sensitivity analysis excluding patient with chronic lung disease did not significantly change the results (data not shown). Age was not found to correlate with poor outcomes, but the cohort was composed of a fairly homogenous group of older patients, limiting this analysis.

The strengths of this study are its multicenter design, thorough case record review by experienced specialists, and its representation of the Israeli population, as it contains approximately 40% of severe COVID-19 patient reported nationally. Nevertheless, some limitations should be noted. The retrospective design might lead to several biases due to inherent differences between patient populations who received varying numbers of vaccine doses. These were adjusted for in the multivariate analyses, but some unknown differences might not have been accounted for. In addition, we excluded patients without valid vaccination records, although these accounted for only 7% of the entire cohort.

## Conclusions

Despite good protection afforded by a 3-dose vaccination schedule against COVID-19, breakthrough infections in vulnerable older populations during an Omicron variant wave resulted in significant morbidity and mortality. Within a population of hospitalized patients with severe/critical COVID-19, recent receipt of a fourth dose resulted in significantly lower probability of death or mechanical ventilation. These findings suggest that administration of a fresh booster dose should be considered for at risk individuals upon an impending new a COVID-19 wave.

## Data Availability

All data produced in the present study are available upon reasonable request to the authors.

